# Sex and Occupation Are Salient Risk Factors for Lateral Ankle Sprain Among Military Tactical Athletes

**DOI:** 10.1101/2020.08.20.20178871

**Authors:** John J. Fraser, Andrew MacGregor, Camille P. Ryans, Mark A. Dreyer, Michael D. Gibboney, Daniel I. Rhon

**Affiliations:** Foot & Ankle Subcommittee, Neuromusculoskeletal Clinical Community Advisory Board, US Navy Bureau of Medicine and Surgery, 7700 Arlington Blvd. Suite 5113. Falls Church, VA 22042-5113, USA; Warfighter Performance Department, Naval Health Research Center, 140 Sylvester Road, San Diego, CA 92106-3251, USA; Medical Modeling, Simulation, and Mission Support Department, Naval Health Research Center, 140 Sylvester Rd, San Diego, CA 92106-3251, USA; Department of Orthopedic Surgery, Naval Hospital Jacksonville, 2104 Massey Ave, Jacksonville, FL, 32228-0148, USA; Podiatry Clinic, Naval Health Clinic New England, 43 Smith Road Newport, RI 02841-1002, USA; Orthopedics Department, Naval Medical Center Camp Lejeune, 100 Brewster Boulevard Camp Lejeune, NC 28547-5008, USA; Physical Performance Service Line, G-3/5/7, US Army Office of the Surgeon General, 5109 Leesburg Pike, Falls Church, VA 22041-3258, USA; Center for the Intrepid, Brooke Army Medical Center, 3551 Roger Brooke Drive Bldg 3600. JBSA Fort Sam Houston, TX 78234-6200, USA

**Keywords:** ankle injuries, military personnel, occupational injuries, sports medicine, public health

## Abstract

**Introduction:** Lateral ankle sprains (LAS) are ubiquitous among tactical athletes and a substantial burden in the military. With the changes in operational demand and the beginning of integration of women into previously closed occupations, an updated assessment of the burden of ankle sprains in the military is warranted.

**Methods:** A population-based epidemiological retrospective cohort study of all service members in the US Armed Forces was performed assessing risk of sex and military occupation on the outcome of LAS incidence. The Defense Medical Epidemiology Database was queried for the number of individuals with ICD-9 diagnosis codes 845.00 (sprain of ankle, unspecified) and 845.02 (calcaneofibular ligament sprain) on their initial encounter from 2006 to 2015. Relative risk (RR) and chi-square statistics were calculated in the assessment of sex and occupational category.

**Results:** A total of 272,970 enlisted males (27.9 per 1000 person-years), 56,732 enlisted females(34.5 per 1000 person-years), 24,534 male officers (12.6 per 1000 person-years), and 6020 female officers (16.4 per 1000 person-years) incurred LAS. Enlisted females in all occupational groups were at significantly higher risk for LAS than their male counterparts (RR 1.09–1.68; *p* < 0.01), except for Engineers (*p =* 0.15). Female officers had consistently higher risk for LAS in all occupational groups (RR 1.10–1.42; *p* < 0.01) compared with male officers, except Ground/Naval Gunfire (*p =* 0.23). Contrasted with Infantry, enlisted members in the Special Operations Forces, Mechanized/Armor, Aviation, Maintenance, and Maritime/Naval Specialties were at lower risk (RR, 0.38–0.93; *p* < 0.01), Artillery, Engineers, and Logistics Specialties were at higher risk (RR 1.04–1.18; *p* < 0.01), and Administration, Intelligence, and Communications were no different (*p* = 0.69). Compared with Ground/Naval Gunfire officers, Aviation officers were at significantly lower risk (RR, 0.75; *p* < 0.01), and Engineers, Maintenance, Administration, Operations/Intelligence, and Logistics officers were at higher risk (RR, 1.08–1.20; *p* < 0.01).

**Conclusion:** Sex and military occupation were salient factors for LAS risk. Colocation of interdisciplinary neuromusculoskeletal specialists to provide targeted preventive interventions should be considered in practice and policy.

**Disclaimer:** The authors are military service members or employees of the U.S. Government. This work was prepared as part of their official duties. Title 17, U.S.C. §105 provides that copyright protection under this title is not available for any work of the U.S. Government. Title 17, U.S.C. §101 defines a U.S. Government work as work prepared by a military service member or employee of the U.S. Government as part of that person’s official duties. The views expressed in this article are those of the authors and do not necessarily reflect the official policy or position of the Department of the Navy, Department of Defense, nor the U.S. Government. The study protocol was approved by the Naval Health Research Center Institutional Review Board in compliance with all applicable Federal regulations governing the protection of human subjects. Research data were derived from an approved Naval Health Research Center Institutional Review Board protocol, number NHRC.2019.0200-NHSR.

**Highlights:**

- Female sex and military occupation were salient factors in risk for LAS.
- These injuries continue to be pervasive among military service members
- Findings likely attribited in part to differences in sex-related musculoskeletal structure and function
- Occupational hazard exposure, physical fitness, and health care access and utilization also likely contributory to LAS risk

## INTRODUCTION

Lateral ankle sprains (LASs) are ubiquitous among military service members[1,2] and the general public.[2] A LAS results from injurious high-velocity inversion and internal rotation moments of the ankle/foot that frequently result in damaged neural, connective, and contractile tissues.[3] While most LASs resolve within 1 year following injury, 40% will progress to develop chronic ankle instability,[4] a condition characterized by persistent perceived or episodic giving way of the ankle that results in activity limitation and participation restriction.[5] Due to the potential impact for degradation of unit readiness resulting from LAS, it is important to identify the burden of LAS in the military and factors that can increase risk for this injury.

Most epidemiological studies of ankle sprain in the military have focused on cadets and midshipmen enrolled in the military academies[6]or in recruit training.[7,8] In the only epidemiological study of ankle sprain in all branches of the US military, Cameron and colleagues[1] investigated the factors of age, sex, and branch of service on LAS rates from 1998 to 2006. This study epoch occurred during the transition from a period of relative peacetime to one of conflict that started following the initiation of Operation Enduring Freedom (OEF) in October 2001 and Operation Iraqi Freedom (OIF) in March 2003, with subsequent drawdowns occurring in 2010 from OIF and 2014 from OEF.

Each branch of service comprises a diverse range of military occupations that have unique requirements for physical performance and encompass diverse environmental hazards. While prior studies have assessed military occupation as a factor in LAS in the US Army,[9,10] it is unclear if these findings are generalizable to other branches of the military. Furthermore, changes in military operations resulting from evolving geopolitics may also influence injury rates. Furthermore, the Secretary of Defense and the Chairman of the Joint Chiefs of Staff released a memorandum in January 2013 rescinding the exclusion of females in previously closed occupation specialties.[11] With the changes in operational demand and the beginning of integration of women into previously closed occupations, an updated assessment of the burden of ankle sprains in the military is warranted. With the increasing integration of women into diverse career fields, it is also important to understand the risk of ankle sprains for both male and female military service members. Hence, the purpose of this epidemiological retrospective cohort study was to assess the risk of LAS in male and female tactical athletes across different military occupations in the US military.

## MATERIALS & METHODS

A population-based epidemiological retrospective cohort study of all service members in the US Armed Forces was performed assessing risk of sex and military occupation on the outcome of LAS incidence. The Defense Medical Epidemiological Database [(DMED), Defense Health Agency, Falls Church, VA, https://bit.ly/DHADMED] was utilized to identify relevant healthcare encounters. This database provides aggregated data for International Classification of Diseases, Ninth Revision (ICD-9) codes and de-identified patient characteristics, including sex, categories of military occupations, and branch of service for all active duty and reserve military service members. The database is HIPAA compliant, does not include any personal identifiable or personal health information, and has been used previously for epidemiological study of ankle injury in the military.[1] This study was approved as non–human-subjects research by the Institutional Review Board at the Naval Health Research Center (NHRC.2019.0200-NHSR).

The database was queried for the number of distinct patients with a primary diagnosis of LAS (ICD-9 code 845.00, sprain of ankle, unspecified site; and 845.02, sprain of calcaneofibular ligament of ankle)[12] on the initial medical encounter in 2006–2015. Patients with repeat visits for the same diagnosis were only counted once in all analyses.

Calculations of cumulative incidence of patients diagnosed with an LAS were conducted for male and female military members, enlisted and officers, in each service branch (Army, Navy, Marine Corps, and Air Force) and occupational category. Relative risk (RR) point estimates and 95% confidence intervals (CIs), risk difference point estimates, attributable risk (AR), number needed to harm (NNH), and chi-square statistics were calculated in the assessment of sex and occupation category, using the group with the lowest rate as the reference group. The level of significance was *p* ≤ .05 for all analyses. RR point estimates were considered statistically significant if CIs did not cross the 1.00 threshold. All calculations were performed using Microsoft Excel for Mac 2016 (Microsoft Corp., Redmond, WA) and a custom epidemiological calculator spreadsheet.[13]

## RESULTS

From 2006 to 2015, 272,970 enlisted males (27.9 per 1000 person-years), 56,732 enlisted females (34.5 per 1000 person-years), 24,534 male officers (12.6 per 1000 person-years), and 6020 female officers (16.4 per 1000 person-years) incurred an LAS. Tables 1 and 2 detail the counts and incidence of LAS for male and female service members in each military occupational category. There was a progressive trend of decreasing LAS rates during the study epoch in all branches, with the exception of male and female Marine Corps officers who experienced increased rates from 2010 onward (Figures 1–4). Table 3 details the findings of sex as a factor in LAS risk within each military occupational category and in each military branch overall. Female enlisted were at significantly higher risk of LAS in all occupational groups compared with their male counterparts (RR, 1.09–1.68; AR, 8.3%-40.4%; NNH, 52–417; *p* < 0.01), with the exception of Engineers (*p =* 0.15). Female officers also had consistently higher risk of LAS in all occupational groups (RR, 1.21–1.42; AR, 17.7%-29.7%; NNH, 183–335; *p* < 0.01) compared with male officers, except Ground/Naval Gunfire (*p =* 0.23). Table 4 details the findings of military occupation as a factor in LAS risk. Enlisted service members in the Special Operation Forces, Mechanized/Armor, Aviation, Maintenance, and Maritime/Naval Specialties were at lower risk and Artillery, Engineers, and Logistics Specialties were at higher risk compared with Infantry. Among officers, Aviation officers were at significantly lower risk and Engineers, Maintenance, Administration, Operations/Intelligence, and Logistics officers were at higher risk than Ground/Naval Gunfire officers. There were no other significant findings.

**Table 1.**
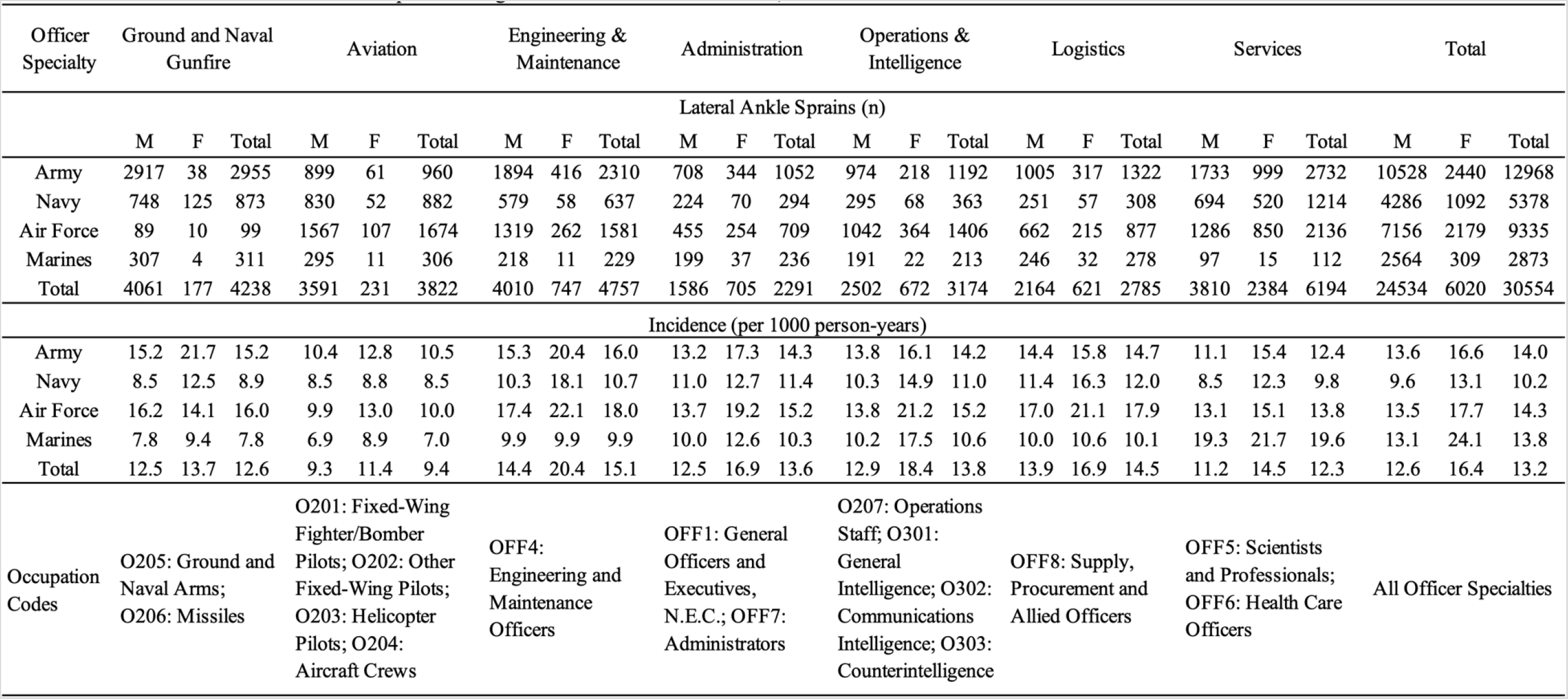
Number and incidance of lateral ankle among officers in the US Armed Forces, 2006–2015

**Table 2.**
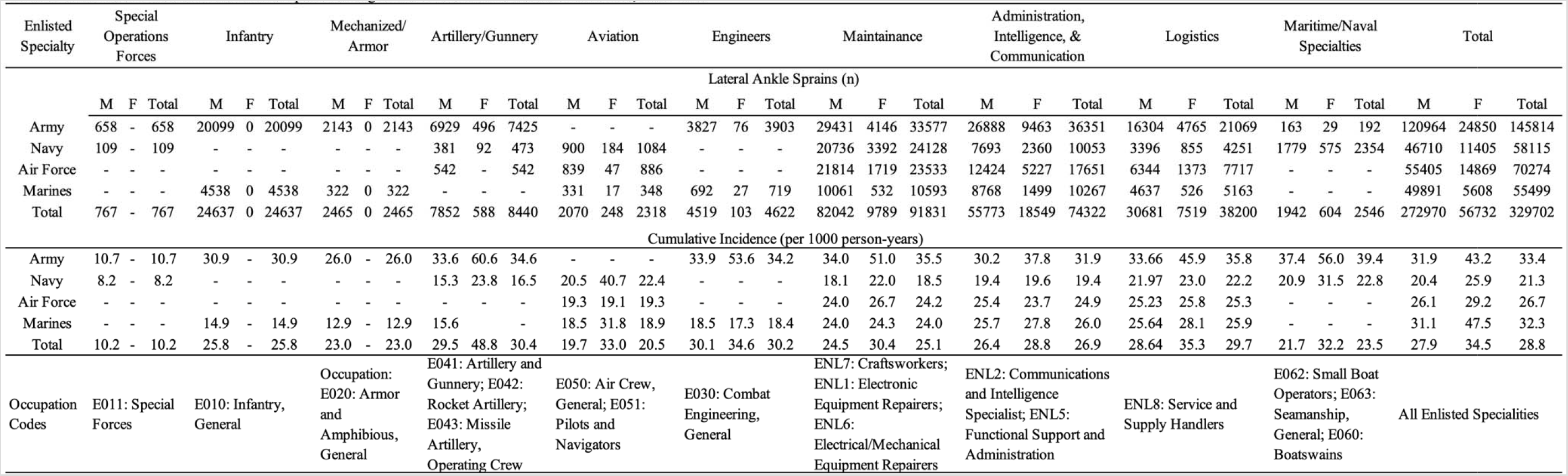
Number and incidance of lateral ankle sprains among elisted mebers in the US Armed Forces, 2006–2015

**Table 3.**
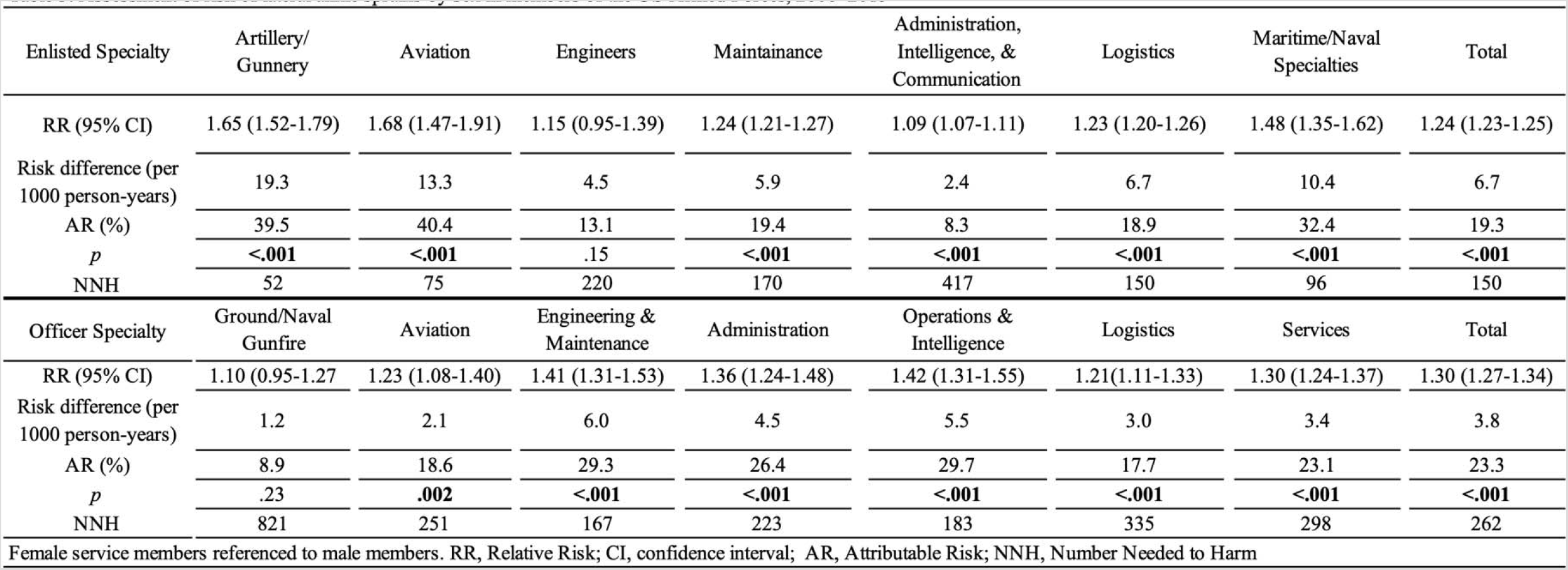
Assesment of risk of lateral ankle sprains by sex in members of the US Armed Forces, 2006–2015

**Table 4.** Assesment of risk of lateral ankle sprains by occupation in members of the US Armed Forces, 2006–2015

| Enlisted Specialty* | Special<br>Operation<br>Forces | Mechanized/<br>Armor | Artillery/<br>Gunnery | Aviation | Engineers | Maintenance | Administration,<br>Intelligence, &<br>Communication | Logistics | Maritime/Naval<br>Specialties |
| --- | --- | --- | --- | --- | --- | --- | --- | --- | --- |
| RR (95% CI) | 0.38 (0.36-0.41) | 0.86 (0.82-0.89) | 1.13 (1.10-1.16) | 0.77 (0.73-0.80) | 1.12 (1.09-1.16) | 0.93 (0.92-0.95) | 1.00 (0.99-1.02) | 1.11 (1.09-1.13) | 0.88 (0.84-0.91) |
| Risk difference (per<br>1000 person-years) | -16.6 | -3.9 | 3.5 | -6.3 | 3.3 | -1.8 | 0.1 | 2.9 | -3.3 |
| AR (%) | -162.2 | -17 | 11.6 | -30.7 | 11.0 | -7.1 | 0.3 | 9.7 | -14.0 |
| *p* | **<.001** | **<.001** | **<.001** | **<.001** | **<.001** | **<.001** | .69 | **<.001** | **<.001** |
| NNH | -60 | -257 | 285 | -159 | 302 | -560 | 13094 | 345 | -303 |
| Officer Specialty† | Aviation<br>Officers | Engineering &<br>Maintenance | Administration | Operations &<br>Intelligence | Logistics | Services |  |  |  |
| RR (95% CI) | 0.75 (0.72-0.78) | 1.20 (1.16-1.26) | 1.08 (1.03-1.14) | 1.10 (1.05-1.15) | 1.15 (1.10-1.21) | 0.98 (0.94-1.02) |  |  |  |
| Risk difference (per<br>1000 person-years) | -3.1 | 2.6 | 1.0 | 1.2 | 1.9 | -0.3 |  |  |  |
| AR (%) | -33.2 | 17.0 | 7.5 | 9.0 | 13.3 | -2.3 |  |  |  |
| *p* | **<.001** | **<.001** | **.003** | **<.001** | **<.001** | .25 |  |  |  |
| NNH | -320 | 389 | 985 | 807 | 521 | -3519 |  |  |  |

\* Contrasted to Enlisted Infantry. † Referenced to Ground and Naval Gunfire Officers. RR, Relative Risk; CI, confidence interval; AR, Attributable Risk; NNH, Number Needed to Harm

**Figure 1.**
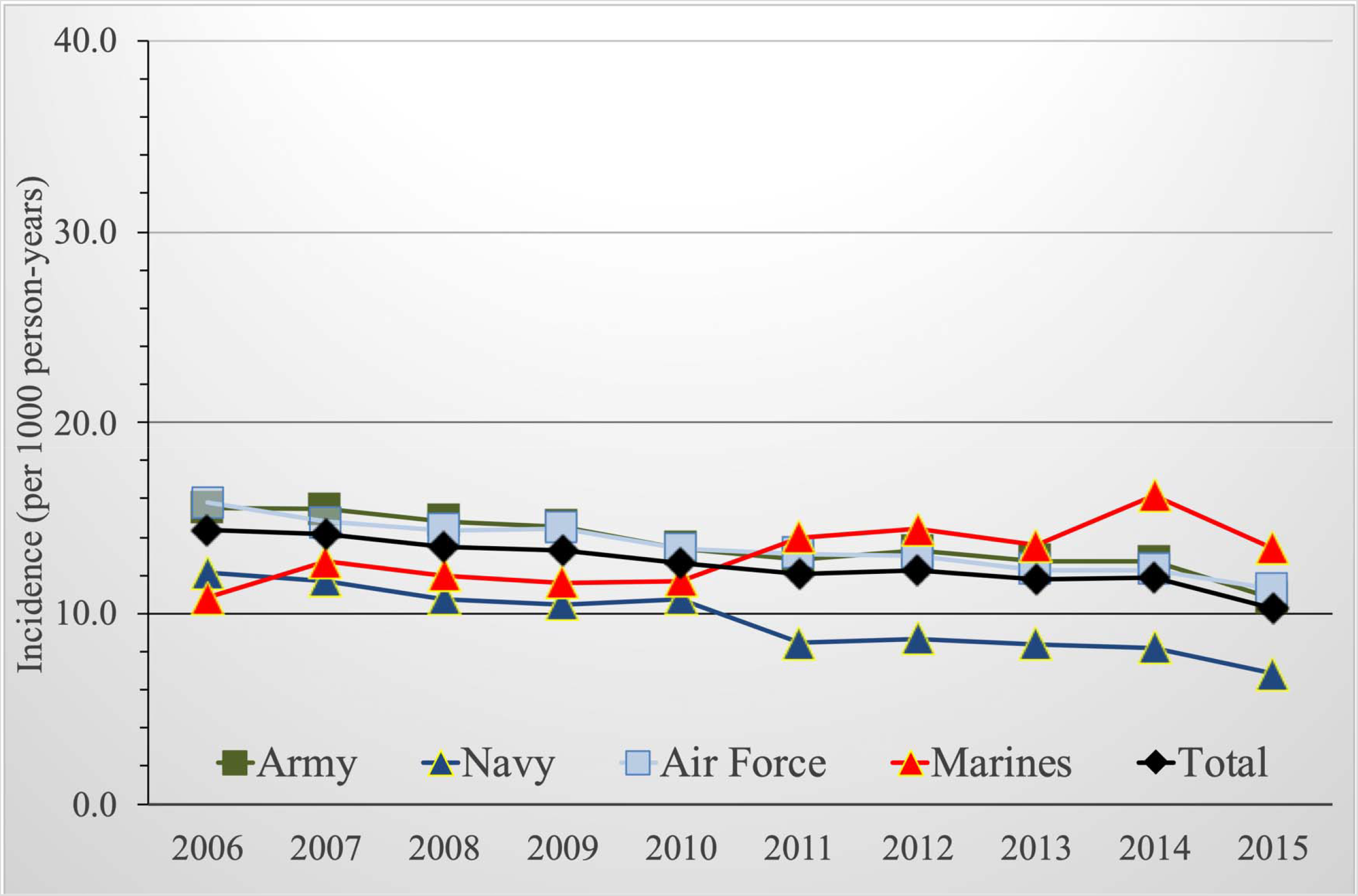
Lateral ankle sprain incidence among male officers, US Armed Forces, 2006–2015.

**Figure 2.**
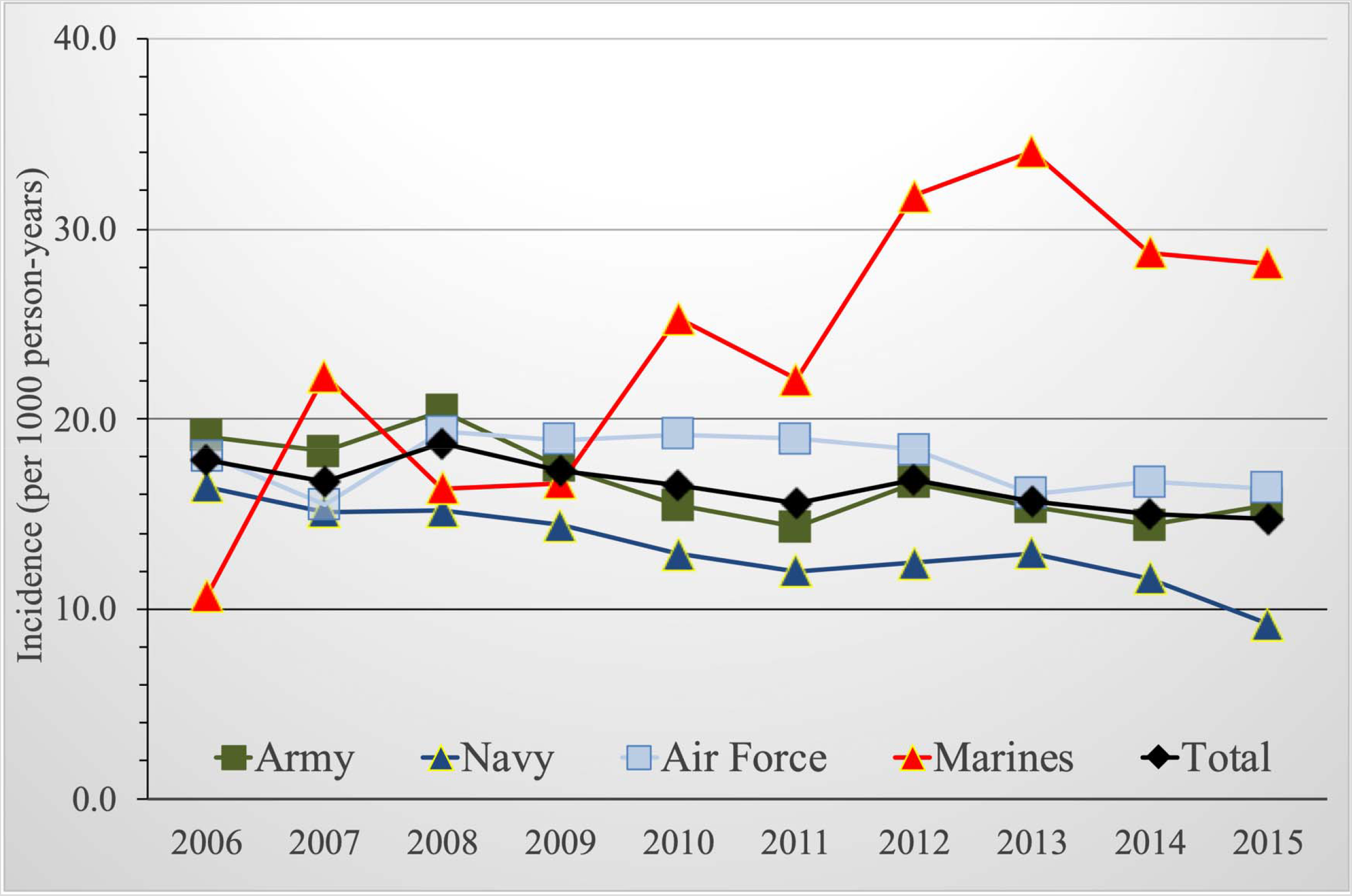
Lateral ankle sprain incidence among female officers, US Armed Forces, 2006–2015.

**Figure 3.**
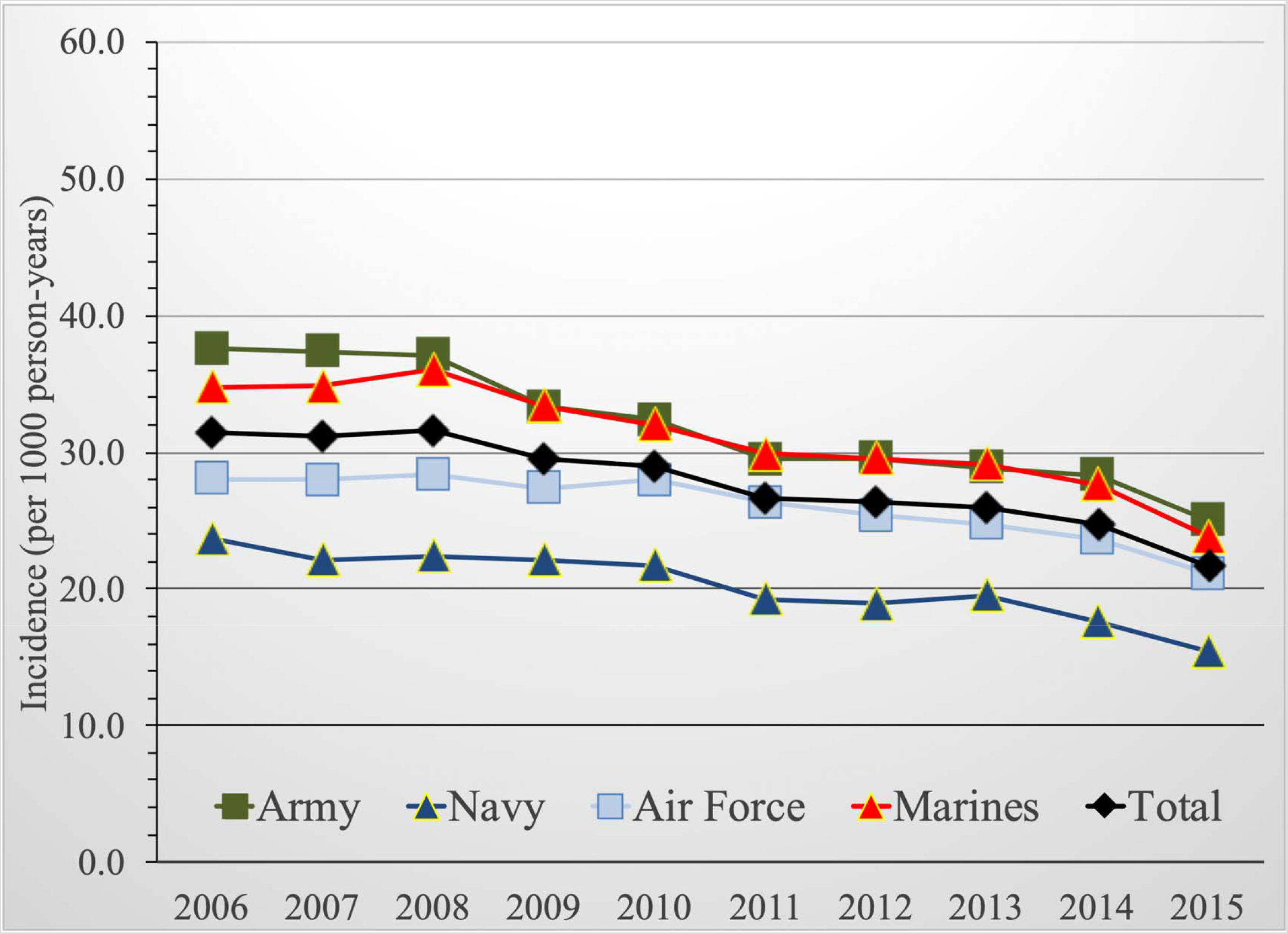
Lateral ankle sprain incidence among enlisted males, US Armed Forces, 2006–2015.

**Figure 4.**
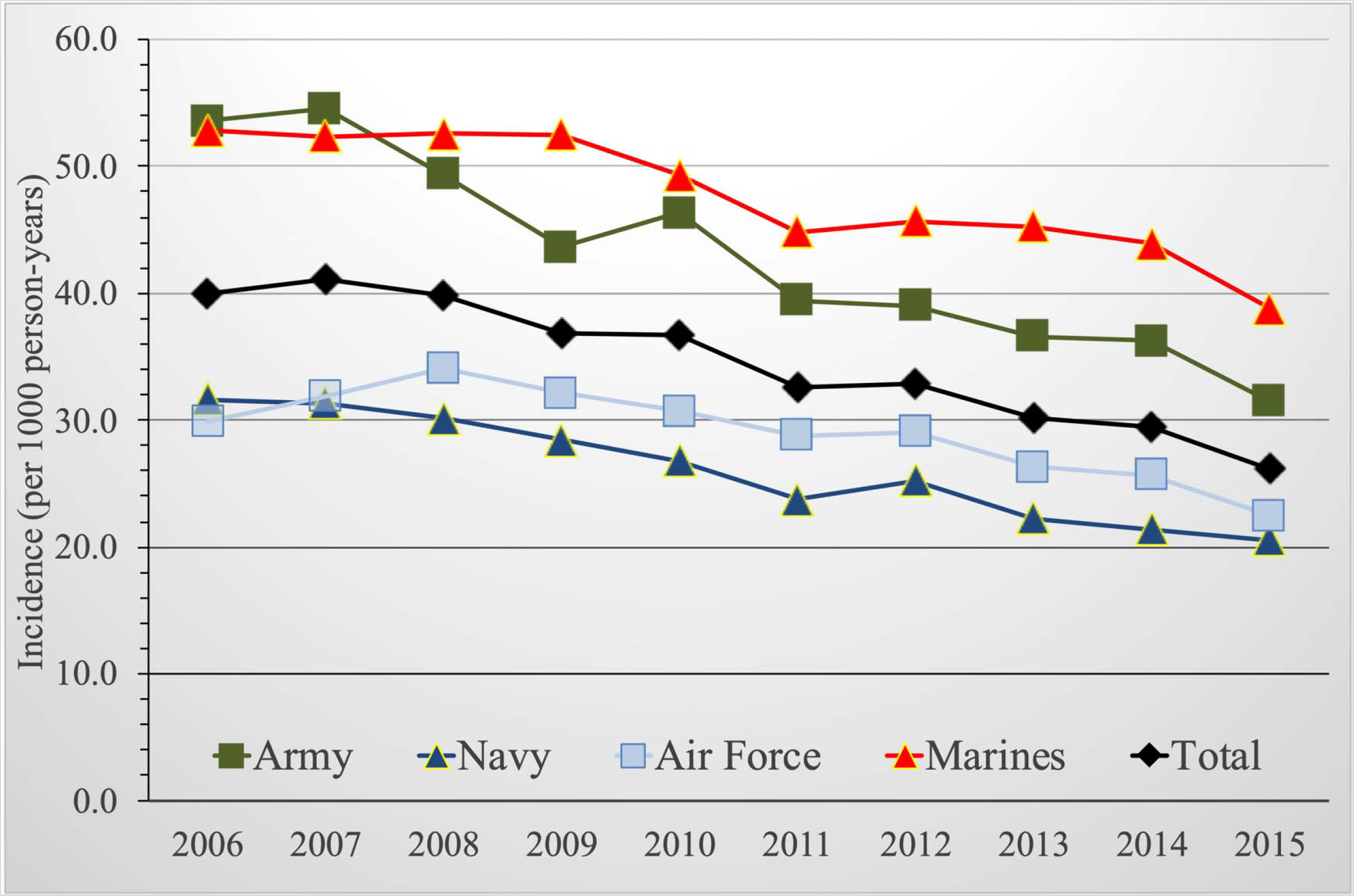
Lateral ankle sprain incidence among enlisted females, US Armed Forces, 2006–2015.

## DISCUSSION

The primary finding was that female military members were at significantly higher risk for incurring an LAS, regardless of military occupation. Additionally, there were varying risks of LAS among the enlisted members and officers that were dependent on military occupation. With the exception of Marine officers, the rates of LAS in the military appear to decrease over the course of the study epoch.

### Injury Rates Over Time

Several factors could explain the overall trend of decreased LAS rates during this study epoch. The beginning of this period coincided with a time of high military operations, with several brief troop surges occurring during OEF (January 2010 to June 2011) and OIF (February 2007 to July 2008).[14] The tempo of military operations substantially slowed during the latter half of the study epoch as a result of redeployment and a relative return to peacetime conditions. We posit that decreased injury exposure resulting from reduced operational demands during training and deployment abroad likely had a protective effect.

There was an increase in LAS rates in male and female Marine officers from 2009 onward. This rise coincided with the implementation of several organization-level changes designed to improve healthcare delivery. First, the implementation of the Marine-Centered Model Home brought additional attention, resources, and accessible medical care to operational units.[15] Second, physical therapists and athletic trainers have been assigned with operational units in proximity to troop billeting and work centers. Reduction in the barriers to care likely increased medical access and capture of LAS diagnoses in these communities. In addition to accessibility, trust in the specialized medical capability to improve functional outcomes and readiness in operational units has likely normalized and encouraged care seeking following injury when it has been traditionally stigmatized.[16] Therefore, this rise in rates could also be partly attributed to improved surveillance and capture of injury in the electronic medical record than an actual increase in the number of injuries.

### Sex

The findings of increased risk of LAS in female service members is consistent with previously published findings. Cameron and colleagues[1] found that female service members were 21% more likely to sustain a LAS than their male counterparts. Waterman et al.[6] similarly found an overall increased risk of LAS in females enrolled at the US Military Academy, a factor that was mediated by athletic participation. While sex-related preferences in physical activity may drive exposure to hazards, physical activity in performance of duty requirements should not differ between sexes within each military occupational group. Sex was a significant risk factor across all but two military occupation groups assessed, with the greatest risk observed among female enlisted in the Artillery/Gunnery, Aviation, and Maritime/Naval specialties and officers in Engineering, Operations, and Intelligence communities.

There are some plausible reasons for why female service members demonstrated increased risk of LAS. Females have been shown to have increased joint laxity compared to males.[17] This is likely a function of joint phenotype,[18] foot morphology,[19] and sex hormonal influences.[20] While military boots worn by male and female service members are of similar design and construction, high-heeled uniform and off-duty footwear choices are likely contributory to increased LAS risk in female service members.[21] Enlisted females in the Artillery/Gunnery, Aviation, and Maritime/Naval communities had AR > 30% compared with their male counterparts. It is difficult to elucidate the reasons why these communities had the highest attributable risk from our data. Female service members are also more likely to seek outpatient health care services than their male counterparts.[22] While sex-related disparities in care seeking could theoretically artificially increase risk among women, our findings are consistent with other studies that used various other methodologies for data collection.[2]

### Occupation

Exposure to hazards that contribute to LAS differ between occupational specialties in the military. In this study, we observed lower rates of injury in the combat arms that primarily employed vehicles (Mechanized/Armor, Aviation, Maritime/Naval Specialties) compared with the Infantry. The Infantry rely on marching and maneuvering over various terrain, in daylight and darkness, to close with and engage a target. Hazards of this task may include stepping on terrain that is inclined, uneven, or contains features such as rocks, boulders, holes, or fallen foliage (ie, twigs, branches, logs) that can contribute to inversion ankle injury. It is likely that sedentary, vehicle-borne, and ground-based occupations that operate on hard, improved, or unimproved surfaces have reduced exposure to hazards that can contribute to LAS.

Differences in physical fitness and body composition among the military occupations are plausible contributions to our findings. Increased adiposity and poor cardiovascular endurance have been identified as risk factors for injury,[23] greater utilization of health care services,[24] and lost duty time[25] in military members. Increased body mass index has also been identified as a risk factor for LAS and prolonged recovery following injury.[26] It is highly plausible that variations in physical activity and adiposity associated with military occupation are contributory to LAS risk. In a study assessing associations of occupation with anthropometry, definitive disparities exist between occupations.[27] This is likely a result of varying occupation-related physical requirements.[28] Physical fitness, to include ankle and foot strength, has been identified as a protective factor for LAS.[29] This may explain in part why Special Operation Forces had a reduced rate of LAS compared with Infantry, despite similar occupational hazards.

This study relied on billed medical encounters, so it is highly plausible that disparity in social determinants of health seeking among different military occupations contributed to our findings.[30] It is estimated that approximately half of all military members who sustain injuries do not seek care from a medical provider.[16] Knowledge, beliefs, attitudes, and values regarding LAS and perceived benefits and risks of seeking medical care among military members may influence care-seeking behaviors. Perceptions that LASs are self-limiting conditions of little consequence, combined with cultural influences among occupational groups, likely reduce care seeking.[30] In some military occupations, such as Special Operators and Aviators, fear of being taken out of operations likely preclude care seeking.[30] Barriers to medical care, such as geographic distance or a high operational tempo, may also limit a service member’s ability to seek treatment.[30] For example, the Army has deployed physical therapists with operational units since the Vietnam War, whereas most Navy and Marine Corps units lose access to this specialized capability once they leave garrison. This may explain why the Navy and Marine Corps have the lowest return-to-duty rates following injury.[31]

### Clinical and Research Implications

As women continue to be integrated into expanding military roles, identification and awareness of risk factors among the general military population and subgroups will be critical in maintaining an operationally ready force. Appropriate preventive practices, such as strengthening, proprioceptive, agility, and flexibility exercise programs, as well as education techniques on bracing, taping, and proper footwear size and fit, are paramount. Collocation of specialized interdisciplinary teams of specialists, such as physical therapists, athletic trainers, and sports medicine physicians with the operational forces for surveillance, prevention, and rehabilitation following injury is paramount both in garrison and when deployed. Future research investigating the knowledge, skills, and abilities required to mitigate injuries such as LAS at each echelon of care is needed. Follow-on study is also required to assess determinants of care seeking and how factors such as adiposity and fitness within the military occupations influence rates of reported injury in the military. Detailed occupational surveys should assess specific work-related exposures and hazards, while longitudinal data collection may be employed to identify barriers to full recovery.

### Strengths and Limitations

Using the DMED allowed for a large, population-based analysis of LASs across all branches of military service. The ability to exclude repeat encounters resulted in an estimate of incidence and gold standard epidemiological measures of risk. Further, the stratification by sex addressed a timely military issue of public health importance given that the full integration of women is currently under way.

There are limitations to this study. The ICD-9 code 845.00 (sprain of ankle, unspecified site) is nonspecific and can refer to medial, lateral, or syndesmotic ankle injury. Many clinicians do not take time to code the specific injury and often just provide the more generic label. However, 845.00 is the most used diagnostic code by clinicians for LAS. While both medial and syndesmotic sprains can also be captured by the 845.00 code, comfort can be taken that that lateral ligament injury represents 90% to 96% of all ankle sprains in the civilian[32] and military sectors.[1] Lastly, these codes do not allow for identification of laterality. A new injury may have occurred on the contralateral side but would not have been counted as a new injury because each individual was only counted once. While we might assume equal on-duty exposure to occupational hazards in both male and female service members, it was not possible to control for sex-related disparity in off-duty activities. Other limitations of this data source include lack of other important factors such as adiposity, physical fitness, history of past or concurrent injury, on- and off-duty exposure, and downstream recurrence rate, which precluded us from performing a more in-depth analysis.

## CONCLUSION

Female sex and military occupation were salient factors in risk for LAS. These injuries continue to be pervasive among military service members and are a substantial threat to operational readiness and tactical performance. These findings are likely attributed to differences in sex-related musculoskeletal structure and function, occupational hazard exposure, physical fitness, and health care access and utilization. Since sex and occupational hazards are salient factors in LAS risk, these factors should be considered when developing and implementing risk-mitigating strategies for training, clinical practice, and policy.

## Data Availability

The data that support the findings of this study are available from the corresponding author upon reasonable request.

## REFERENCES

[1] Cameron KL, Owens BD, DeBerardino TM. Incidence of ankle sprains among active-duty members of the United States Armed Services from 1998 through 2006. J Athl Train 2010;45:29–38. https://doi.org/10.4085/1062-6050-45.1.29.

[2] Doherty C, Delahunt E, Caulfield B, Hertel J, Ryan J, Bleakley C. The incidence and prevalence of ankle sprain injury: a systematic review and meta-analysis of prospective epidemiological studies. Sports Med 2014;44:123–40. https://doi.org/10.1007/s40279-013-0102-5.

[3] Fraser JJ, Hertel J. Joint Mobility & Stability Strategies for the Ankle. 292 Neurol. Orthop., Alexandria, VA: Academy of Orthopaedic Physical Therapy, APTA, Inc.; 2019

[4] Doherty C, Bleakley C, Hertel J, Caulfield B, Ryan J, Delahunt E. Recovery from a first-time lateral ankle sprain and the predictors of chronic ankle instability: a prospective cohort analysis. Am J Sports Med 2016;44:995–1003. https://doi.org/10.1177/0363546516628870.

[5] Gribble PA, Delahunt E, Bleakley C, Caulfield B, Docherty CL, Fourchet F, et al. Selection criteria for patients with chronic ankle instability in controlled research: a position statement of the International Ankle Consortium. J Orthop Sports Phys Ther 2013;43:585–91. https://doi.org/10.2519/jospt.2013.0303.

[6] Waterman BR, Belmont PJ, Cameron KL, DeBerardino TM, Owens BD. Epidemiology of ankle sprain at the United States Military Academy. Am J Sports Med 2010;38:797–803. https://doi.org/10.1177/0363546509350757.

[7] Almeida SA, Williams KM, Shaffer RA, Brodine SK. Epidemiological patterns of musculoskeletal injuries and physical training: Med Sci Sports Exerc 1999;31:1176–82. https://doi.org/10.1097/00005768-199908000-00015.

[8] Linenger J, Shwayhat A. Epidemiology of podiatric injuries in US Marine recruits undergoing basic training. J Am Podiatr Med Assoc 1992;82:269–71. https://doi.org/10.7547/87507315-82-5-269.

[9] Bulathsinhala L, Hill OT, Scofield DE, Haley TF, Kardouni JR. Epidemiology of ankle sprains and the risk of separation from service in US Army soldiers. J Orthop Sports Phys Ther 2015;45:477–84. https://doi.org/10.2519/jospt.2015.5733.

[10] Teyhen DS, Goffar SL, Shaffer SW, Kiesel K, Butler RJ, Tedaldi A-M, et al. Incidence of musculoskeletal injury in US Army unit types: a prospective cohort study. J Orthop Sports Phys Ther 2018;48:749–57. https://doi.org/10.2519/jospt.2018.7979.

[11] Dempsey ME, Panetta LE. Elimination of the 1994 Direct Ground Combat Definition and Assignment Rule. Memorandum for Secretaries of the Military Departments Acting Under Secretary of Defense for Personnel and Readiness, and Chiefs of the Military Services. Washington, DC: Joint Chiefs of Staff; 2013

[12] Feger MA, Glaviano NR, Donovan L, Hart JM, Saliba SA, Park JS, et al. Current Trends in the Management of Lateral Ankle Sprain in the United States. Clin J Sport Med Off J Can Acad Sport Med 2016;27:1. https://doi.org/10.1097/JSM.0000000000000321.

[13] LaMorte WW. Epidemiology/Biostatistics Tools [Excel workbook]. n.d.

[14] Peters HM, Plagakis S. Department of Defense Contractor and Troop Levels in Afghanistan and Iraq: 2007–2018. Washington, DC: Congressional Research Service; 2019

[15] Nathan ML. The Patient-Centered Medical Home in the transformation from healthcare to health. Mil Med 2013;178:126–7. https://doi.org/10.7205/MILMED-D-12-00467.

[16] Smith L, Westrick R, Sauers S, Cooper A, Scofield D, Claro P, et al. Underreporting of musculoskeletal injuries in the US Army. Sports Health 2016;8:507–13. https://doi.org/10.1177/1941738116670873.

[17] Wilkerson RD, Mason MA. Difference in men’s and women’s mean ankle ligamentous laxity. Iowa Orthop J 2000;20:46–8.

[18] Kim SK, Kleimeyer JP, Ahmed MA, Avins AL, Fredericson M, Dragoo JL, et al. Two genetic loci associated with ankle injury. PLoS ONE 2017;12:e0185355. https://doi.org/10.1371/journal.pone.0185355.

[19] Segal NA, Boyer ER, Teran-Yengle P, Glass NA, Hillstrom HJ, Yack HJ. Pregnancy leads to lasting changes in foot structure. Am J Phys Med Rehabil 2013;92:232–40. https://doi.org/10.1097/PHM.0b013e31827443a9.

[20] Chidi-Ogbolu N, Baar K. Effect of estrogen on musculoskeletal performance and injury. Front Physiol 2019;9:1834. https://doi.org/10.3389/fphys.2018.01834.

[21] Williams CM, Haines TP. An exploration of emergency department presentations related to high heel footwear in Victoria, Australia, 2006–2010. J Foot Ankle Res 20147:4. https://doi.org/10.1186/1757-1146-7-4.

[22] Duggal M, Goulet JL, Womack J, Gordon K, Mattocks K, Haskell SG, et al. Comparison of outpatient health care utilization among returning women and men Veterans from Afghanistan and Iraq. BMC Health Serv Res 2010;10:175.

[23] Anderson MK, Grier T, Canham-Chervak M, Bushman TT, Jones BH. Occupation and other risk factors for injury among enlisted U.S. Army Soldiers. Public Health 2015;129:531–8. https://doi.org/10.1016/j.puhe.2015.02.003.

[24] Shiozawa B, Madsen C, Banaag A, Patel A, Koehlmoos T. Body mass index effect on health service utilization among active duty male United States Army soldiers. Mil Med 2019. https://doi.org/10.1093/milmed/usz032.

[25] Kyröläinen H, Häkkinen K, Kautiainen H, Santtila M, Pihlainen K, Häkkinen A. Physical fitness, BMI and sickness absence in male military personnel. Occup Med 2008;58:251–6. https://doi.org/10.1093/occmed/kqn010.

[26] Bielska IA, Brison R, Brouwer B, Janssen I, Johnson AP, Day AG, et al. Is recovery from ankle sprains negatively affected by obesity? Ann Phys Rehabil Med 2019;62:8–13. https://doi.org/10.1016/j.rehab.2018.08.006.

[27] Gu JK, Charles LE, Bang KM, Ma CC, Andrew ME, Violanti JM, et al. Prevalence of Obesity by Occupation Among US Workers. J Occup Environ Med Am Coll Occup Environ Med 2014;56:516–28. https://doi.org/10.1097/JOM.0000000000000133.

[28] Church TS, Thomas DM, Tudor-Locke C, Katzmarzyk PT, Earnest CP, Rodarte RQ, et al. Trends over 5 decades in U.S. occupation-related physical activity and their associations with obesity. PLoS ONE 2011;6:e19657. https://doi.org/10.1371/journal.pone.0019657.

[29] Delahunt E, Remus A. Risk factors for lateral ankle sprains and chronic ankle instability. J Athl Train 2019;54:611–6. https://doi.org/10.4085/1062-6050-44-18.

[30] Fraser JJ, Schmied E, Rosenthal MD, Davenport TE. Physical Therapy as a Force Multiplier: Population Health Perspectives to Address Short-Term Readiness and Long-Term Health of Military Service Members. Cardiopulm Phys Ther J 2020;31:22–8. https://doi.org/10.1097/CPT.0000000000000129.

[31] Cohen SP, Brown C, Kurihara C, Plunkett A, Nguyen C, Strassels SA. Diagnoses and factors associated with medical evacuation and return to duty for service members participating in Operation Iraqi Freedom or Operation Enduring Freedom: a prospective cohort study. The Lancet 2010;375:301–9. https://doi.org/10.1016/S0140-6736(09)61797-9.

[32] Shah S, Thomas AC, Noone JM, Blanchette CM, Wikstrom EA. Incidence and cost of ankle sprains in United States emergency departments. Sports Health 2016;8:547–52. https://doi.org/10.1177/1941738116659639.

